# Obesity is an Independent Risk Factor for In-Hospital Mortality among Patients Admitted for Heart Failure: A National Study

**DOI:** 10.1101/2023.05.12.23289919

**Authors:** Liyun Liu, Ye Zhu, Olamide Oyenubi, M. Hassan Murad, Stephen Jesmajian

**Author notes:** <u>Corresponding Author</u>: Liyun Liu, MD, PhD, Department of Internal Medicine, Montefiore New Rochelle Hospital, 16 Guion Place, New Rochelle, NY 10801.

## Abstract

**Background:** Obesity is a major risk factor for developing chronic conditions such as cardiovascular disease and diabetes. However, the phenomenon of “obesity paradox” has been reported over the past two decades which makes the relationship between BMI and inpatient mortality unclear.

**Methods:** This study identified adult patients (aged 18 years or older) who were admitted to hospitals for the primary cause of heart failure during 2017-2019 from the Healthcare Cost and Utilization Project (HCUP) National Inpatient Sample (NIS) database. Baseline characteristics (i.e., weight status, age, gender, race, Elixhauser score) at the time of admission and the comorbidities were collected. Associations between weight status and in-hospital mortality were examined using logistic regression models that adjusted for individual comorbidities and global risk measures. The likelihood of patients developing each disease comorbidity under different obesity states was examined using logistic regression and the odds were compared across all the disease comorbidities.

**Results:** The study identified 204,970 hospital admissions with 4,290 (2.1%) deaths during the hospitalization and 200,680 (97.9%) live discharges. Analysis that did not adjust for individual comorbidities demonstrated the paradox. However, when adjusting for individual comorbidities and global risk measures and compared to the normal-weight patients, those who had higher BMI had an increased risk for in-hospital mortality. BMI of 35-39.9 group had a 26.5% higher likelihood of in-hospital mortality (OR=1.265, 95% CI: 1.066 - 1.503); BMI of 40-69.9 groups was 61.0% to 83.8% higher odds to die in hospital (OR ranged from 1.610 to 1.838, 95% CI varied); patients with a BMI of 70 and above had higher odds of in-hospital mortality (OR=3.144, 95%CI: 2.351 - 4.203).

**Conclusion:** Obesity is an independent risk factor for in-hospital mortality among patients who were admitted for heart failure. Adjustment for individual comorbidities resolves the obesity paradox. Patients with obesity have a different spectrum of diseases compared to non-obese patients, which may lead to the obesity paradox and bias in the inpatient outcome evaluation.

**Clinical Perspective:** *What is new?:*

- Obesity is an independent risk factor for in-hospital mortality.
- In-hospital mortality increases with the increase in BMI among patients with obesity.
- The obesity-paradox in in-hospital mortality may be due to the fact that obese patients carry a different spectrum of diseases compared to normal-weight patients, which may bias the health outcomes.

*What are the clinical implications?:*

- Maintaining a healthy weight is important in the disease management of patients with congestive heart failure.

**Data Availability Statement:** The data that support the findings of this study are available from Health Care Cost and Utilization Project (H-CUP) at AHRQ. Restrictions apply to the availability of these data, which were used under approval for this study. Data are available https://hcup-us.ahrq.gov/db/nation/nis/nisdbdocumentation.jsp with the permission of AHRQ.

## Introduction

Obesity has been a challenging condition worldwide. It is defined by the Body Mass Index (BMI) in adults, which is calculated from height and weight, which theoretically should be adjusted for muscle mass and waist circumference. Despite current progress in medicine, the prevalence of obesity keeps trending up and affecting over 40 % of the US population as per CDC 2020.^1^

Obesity has been found to increase overall death in the US by 20%-83%, and warrants careful patient management.^2^ Evidence has shown that obesity is a major risk factor for developing conditions such as cardiovascular disease, diabetes, muscular disorders, and certain cancers,^2,3^ as well as increased all-cause mortality ^4^. Data were consistent across worldwide, with the lowest mortality hazard ratio in the 20-25 kg/m2 group, following a 5-year follow-up.

However, the relationship between BMI levels and inpatient outcomes has not been fully established. Numerous studies have also reported a phenomenon known as the “obesity paradox”, in which increased BMI was associated with lower mortality in various conditions.^5-8^ For example, a large cohort study concluded that in advanced heart failure, increased BMI was associated with survival benefits.^5^ The mechanisms and causality of the obesity paradox have not been fully understood.

Similarly, favorable effects of BMI were also observed in other conditions, such as chronic obstructive pulmonary disease, pneumonia, and liver cirrhosis. This was not consistent with previous findings, and no convincing explanations could prove this phenomenon. Not surprisingly, conflicting reports were published suggesting the conclusion of the obesity paradox may require a second thought. Banack used renal cell carcinoma as an example and hypothesized that collider stratification bias as well as failure to acknowledge illness-related weight loss, could potentially reverse the outcome of an effect (e.g, obesity).^9^ Borisov used a randomized controlled trial and found no paradox in community-acquired pneumonia.^10^ Worth mentioning, neither adverse nor beneficial effects of obesity on clinical stability were observed in this study. Some studies were able to identify the association between higher BMI with mortality and increased length of hospital stay, although the down-trending of mortality with BMI> 40 was not expected.^11^

To further evaluate the relationship between BMI and mortality, especially in acute settings, we designed this study using the nationwide inpatient database. All patients admitted due to heart failure from 2017 to 2019 were included in the current study. While the majority of the previous studies adjusted for mortalities using global risk measures, ^5,9,10^ we used a unique approach, which adjusted for both individual comorbidities separately and global risk measures to examine the effect of different BMI categories on in-hospital mortality. It may also guide policy considerations for including a patient’s weight into outcome prediction.

## Methods

### Data and Study Population

The study sample was selected from the Healthcare Cost and Utilization Project (HCUP) National Inpatient Sample (NIS) database. This database is a national-level inpatient database that includes patients drawn from all hospitals based on a random sampling weighted by hospital location, ownership, teaching status, bed size, diagnosis of hospital stay, and month of admission.^12^ This database provided the patient characteristics, disease diagnosis, inpatient procedures, hospital characteristics, and costs. In this study, we identified adult patients (aged 18 years or older) who were admitted to hospitals for the primary cause of congestive heart failure during 2017-2019. Patient’s age, gender, race, length of stay, and year of admission were collected.

ICD-10 codes were used to identify study variables (Appendix eTable 1). For each hospital stay, the first diagnosis (Dx1) was used as the cause of admission and those who were admitted for heart failure were included in this study. Weight status was identified using the ICD10 codes, which were categorized into 10 BMI groups (< 19.9, 20-24.9, 25-29.9, 30-34.9, 35-39.9, 40-44.9, 45-49.9, 50-59.9, 60-69.9, and >= 70).

The rest of the diagnoses (Dx2 – Dx40) were recorded as the disease comorbidities present at the time of hospitalization. Disease comorbidities used in the calculation of the Elixhauser score, which has been shown to be the most associated with in-hospital mortality, were included as comorbidities of interest.^13^

### Statistical Analysis and Regression Models

Descriptive analysis (unadjusted analysis) was performed to provide information for the baseline characteristics of the patients (i.e., weight status, age, gender, race, Elixhauser score,) at the time of admission and the comorbidities at presentation. Baseline differences were compared between patients who were discharged alive and patients who died in the hospital.

Logistic regression was used to calculate the associations between weight status and in-hospital mortality, with odds ratios reported. Models controlled for age, gender, race, length of stay, year of admission, Elixhauser scores, and individual comorbidities. NIS discharge weights were applied in the estimation.

We defined p-value <0.05 as statistically significant and 95% confidence intervals (95% CI) were reported to estimate uncertainty. Analyses were performed using Stata/SE 16.1 (StataCorp LP, College Station, TX).

## Results

### Descriptive and unadjusted Analysis

The study identified 204,970 hospital admissions with 4,290 (2.1%) deaths during hospitalization and 200,680 (97.9%) live discharges. Table 1 listed the baseline characteristics of the study population, and the comparison between in-hospital death and live discharges by each characteristic. Weight status was significantly different between patients who died and those who were discharged alive. Overall, underweight patients had the highest death rate (5.9%), while normal-weight patients had a mortality rate of 4.3%, overweight patients had a mortality rate of 3.0%, and obese patients had a mortality rate of 1.6%. Among obese patients, those with BMI 35-39.9 and BMI 60-69.9 had the lowest mortality rate (1.4%), while other groups had up to 2.1% mortality rate (p<0.0001).

**Table 1.**
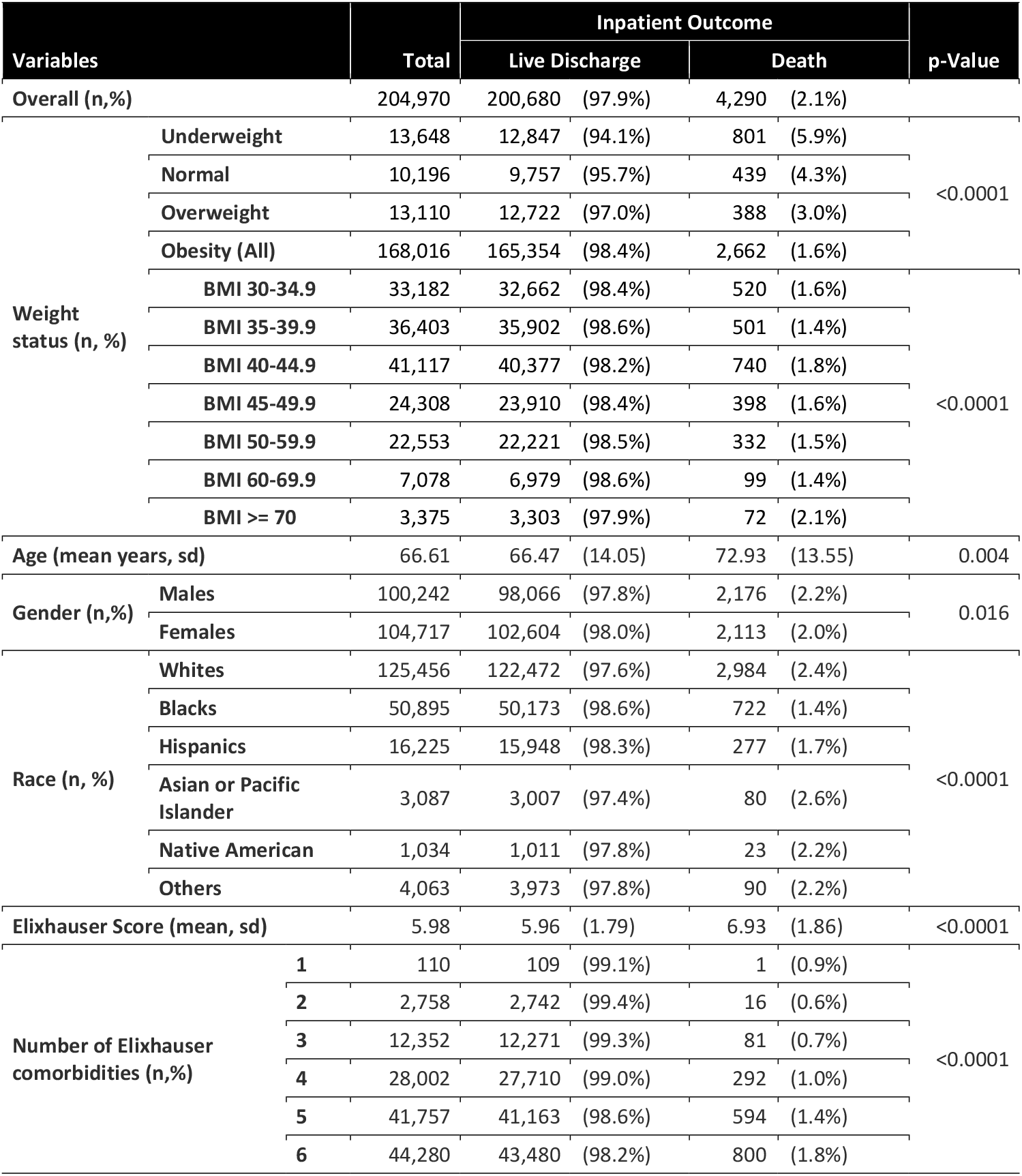
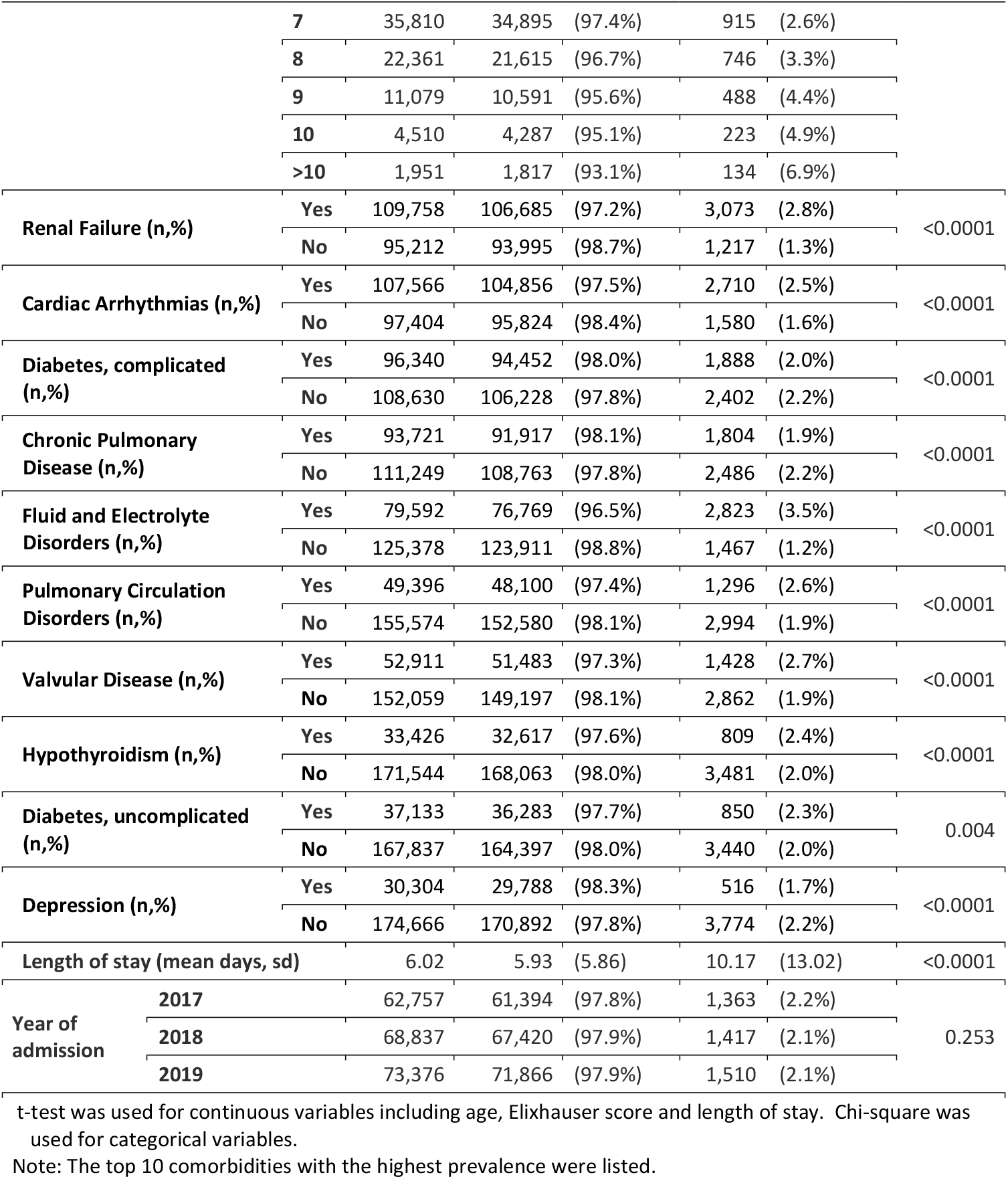
Baseline characteristics of the study cohort (descriptive and unadjusted)

Patients who died in the hospital were older than those who were discharged alive (72.93 vs. 66.47 years, p<0.0001), and men had a higher mortality rate than women (2.2% vs. 2.0%, p=0.016). Patients who died in the hospital had higher Elixhauser scores (6.93 vs. 5.96, p<0.0001). Mortality rates increased from 0.9 % for patients with one comorbidity to 6.9% for patients with more than 10 comorbidities. Patients with comorbidities such as renal failure, cardiac arrhythmias, etc, had significantly higher in-hospital mortality rates than those without such conditions, while patients with comorbidities such as depression had lower in-hospital deaths than those without depression. The patients who died in the hospital had a longer hospital stay than those who were discharged alive (10.17 vs. 5.93 days, p<0.0001). The overall prevalences of disease comorbidities are listed in Appendix eTable 2.

### Multivariate Analysis

The regression results suggested that obesity was associated with in-hospital mortality (Table 2). Compared with the normal weight: overweight patients had an 18% higher likelihood of dying in hospital (OR=1.182, 95% CI: 1.017 -1.373); patients with a BMI of 35-39.9 had a 26.5% higher likelihood of in-hospital mortality (OR=1.265, 95% CI: 1.066 -1.503); patients with a BMI of 40-69.9 were 61.0% to 83.8% higher odds to die in hospital than normal patients (OR ranged from 1.610 to 1.838, 95% CI varied); patients with a BMI of 70 and above had higher odds of in-hospital mortality than normal patients (OR=3.144, 95%CI: 2.351 -4.203).

**Table 2.** The associations between the patient characteristics and in-hospital mortality adjusted for disease comorbidities.

| Patient Characteristics |  | Odds Ratio | 95% CI |
| --- | --- | --- | --- |
| Weight status | Normal |  | Reference |
|  | Underweight | 1.409* | (1.238 - 1.603) |
|  | Overweight | 1.182* | (1.014 - 1.377) |
|  | Obese Group 1 (BMI 30-34.9) | 1.165 | (0.985 - 1.377) |
|  | Obese Group 2 (BMI 35-39.9) | 1.265* | (1.060 - 1.511) |
|  | Obese Group3 (BMI 40-44.9) | 1.610* | (1.355 - 1.914) |
|  | Obese Group 4(BMI 45-49.9) | 1.662* | (1.377 - 2.006) |
|  | Obese Group 5 (BMI 50-59.9) | 1.622* | (1.331 - 1.976) |
|  | Obese Group 6 (BMI 60-69.9) | 1.838* | (1.414 - 2.390) |
|  | Obese Group 7 (BMI >= 70) | 3.144* | (2.345 - 4.214) |
| Age |  | 1.024* | (1.020 - 1.027) |
| Gender | Males |  | Reference |
|  | Females | 0.854* | (0.798 - 0.915) |
| Race | Whites |  | Reference |
|  | Blacks | 0.763* | (0.698 - 0.834) |
|  | Hispanics | 0.877* | (0.769 - 1.000) |
|  | Asian or Pacific Islander | 0.797 | (0.626 - 1.016) |
|  | Native American | 1.161 | (0.747 - 1.802) |
|  | Others | 0.977 | (0.779 - 1.224) |
| Length of Stay |  | 1.022* | (1.019 - 1.026) |
| Neurological Disorders Other Than Paralysis |  | 6.428* | (5.608 - 7.369) |
| Liver Disease |  | 4.775* | (4.099 - 5.564) |
| Fluid and Electrolyte Disorders |  | 4.640* | (4.063 - 5.300) |
| Metastatic Cancer |  | 3.902* | (2.959 - 5.145) |
| Coagulopathy |  | 3.650* | (3.156 - 4.221) |
| Paralysis |  | 3.611* | (2.584 - 5.048) |
| Cardiac Arrhythmias |  | 3.368* | (2.941 - 3.857) |
| Lymphoma |  | 2.844* | (2.146 - 3.771) |
| Recent Weight Loss |  | 2.810* | (2.465 - 3.203) |
| Hypertension, uncomplicated |  | 2.760* | (2.125 - 3.585) |
| Year of Admission | 2017 |  | Reference |
|  | 2018 | 0.926 | (0.855 - 1.003) |
|  | 2019 | 0.899* | (0.831 - 0.972) |

\* p<0.05
Comorbidities with the top 10 odds ratios were listed.

Among the covariates, all 29 comorbidities were significantly associated with higher odds of in-hospital mortality. Neurological disorders other than paralysis, (e.g., epilepsy, demyelination of the central nervous system, Parkinson’s disease) were associated with the highest odds of in-hospital mortality (OR=6.428, 95%CI: 5.649 -7.315). Liver disease was associated with 3 times higher in-hospital mortality (OR=4.775, 95%CI: 4.126 -5.527). Metastatic cancer was associated with an approximately 3-fold increase in in-hospital mortality (OR=4.640, 95%CI: 4.074 -5.285). Coagulopathy was associated with 2.65 times higher mortality (OR=3.650, 95%CI: 3.177 -4.192).

Other covariates had less impact on the in-hospital mortality comparable to that of disease comorbidities. The odds of dying in hospital increased by 2.4% with each year of age increase (OR=1.024, 95% CI: 1.020 -1.027). Women were 14.6% less likely to die in hospital than men (OR=0.854, 95%CI: 0.798 -0.914). Compared to white patients, black and Hispanic patients were less likely to die in the hospital after controlling for all other factors (OR=0.763, 95% CI: 0.699 -0.834 and OR=0.877, 95% CI: 0.770 -0.998). Other races (i.e., Asian or Pacific islanders, native American, or others) did not differ from white patients. Each one-day increase in length of stay, the in-hospital mortality was associated with a 2% increase in in-hospital mortality (OR=1.022, 95% CI: 1.020 -1.025). Compared to admissions in 2017, hospitalizations in 2019 were associated with a lower likelihood of in-hospital mortality (OR=0.899, 95% CI: 0.832 -0.971), while 2018 was not significantly different (OR=0.926, 95% CI: 0.856 -1.002).

The prevalence of each disease comorbidity in the four major weight groups was calculated and listed by the rate among obesity patients (Figure 1A). Figure 1B illustrated the paired odds ratio of each disease comorbidity on in-hospital mortality among the patients with that specific disease compared to patients without the disease. Renal failure was ranked as the comorbidity with the highest rate (52.4%), followed by cardiac arrhythmias (52.2%), complicated diabetes (49.7%), chronic pulmonary diseases (48.3%) and fluid and electrolyte disorders (37.3%). For patients with underweight, normal weight and overweight, the top five comorbidities were recent weight loss (39.2%, 28.4%, and 14.9% respectively), tumor without metastasis (13.7%, 11.8%, and 10.6% respectively), metastatic cancer (15.2%, 15.5%, and 12.0% respectively), lymphoma (14.5%, 12.2%, and 11.5% respectively), and AIDS (20.2%, 12.4%, and 7.5% respectively). The corresponding odds of in-hospital mortality (from the multivariate logistic regression) were listed in Figure 1B.

**Figure 1.**
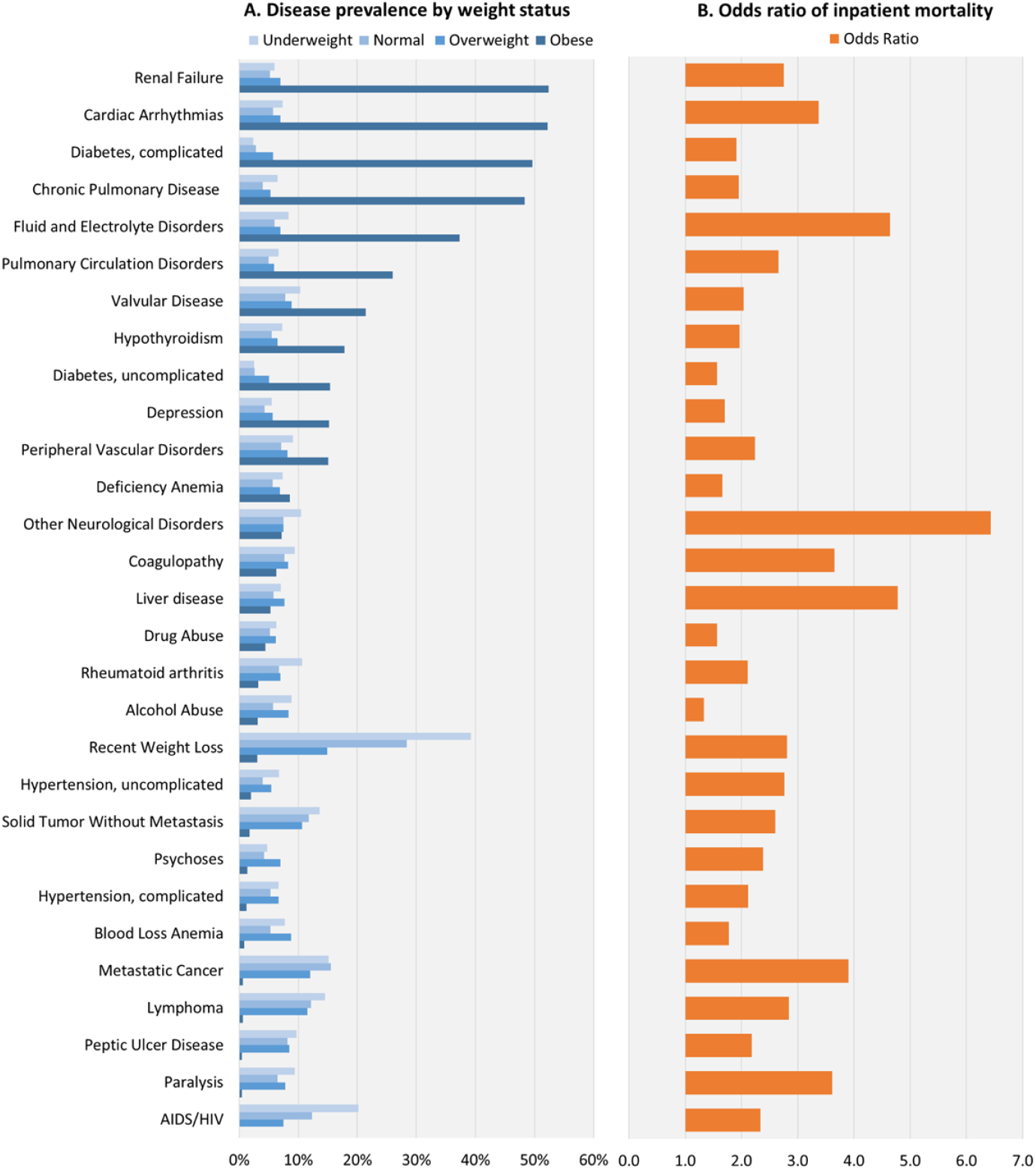
The prevalence rate among hospitalized patients admitted for acute heart failure by weight status and the odds ratio of in-hospital mortality for each disease comorbidity.

## Discussion

This study found that obesity is an independent risk factor for in-hospital mortality in patients admitted to the hospital with heart failure as the primary diagnosis. When other variables were held constant, the odds of in-hospital mortality for overweight and obese patients increased with the increase in BMI.

Patients with obesity presented with various disease comorbidities which lead to different inpatient mortality. Neurological disorders other than paralysis (e.g., Parkinson’s disease, epilepsy, degeneration of nervous system, etc.), liver disease, fluid, and electrolyte disorders bore the highest risk of in-hospital mortality, but they were less likely to be seen in obese patients than in normal weight patients when they were admitted to the hospital.

In our study, we observed the phenomenon that patients with obesity were more likely to have lower in-hospital mortality rates when using the method of descriptive and unadjusted analysis Table 1). Furthermore, we also found that when individual disease morbidity was not controlled, the obesity paradox could also be present (Appendix eTable 3). This observation is consistent with other studies where the obesity paradox was observed.^8,14-16^ However, when individual comorbidities were controlled for separately, in addition to a global risk measure (comorbidity scores such as Elixhauser scores), the paradoxical association no longer exists. This suggested the paradox may be caused by the different comorbidity profiles developed in obese patients.

To further uncover the underlying reasons, we compared disease comorbidities among these different weight statuses of patients. We found that patients with obesity had a higher prevalence of comorbidities that had relatively lower inpatient mortality, which may explain why obese patients had fewer in-hospital deaths in numbers than normal-weight patients as noted in the unadjusted analysis ( Table 1). When compared across different weight statuses, patients with obesity had lower Elixhauser scores than normal-weight patients (5.96 vs. 6.12, eTable 4). It might be related to illness-related weight loss, which was raised as a hypothesis for the phenomenon of the obesity paradox.^9^

Therefore, it is important to adjust for disease comorbidities to separate the effect of obesity from the effect of disease comorbidities (Table 2). For example, patients with other neurological diseases (neurology disease other than paralysis) had much higher odds of in-hospital death than a patient with uncomplicated diabetes (OR=6.428 vs. OR=1.564, Figure 1). If an obese patient had uncomplicated diabetes and a normal-weight patient had neurological disease other than paralysis, we may observe that the obese (and diabetic) patient was discharged alive while the normal-weight patient (with other neurological disease) died in the hospital. This bias was caused by the unbalanced risks between the obese patient and the normal-weight patient. Therefore, when we compare these two patients, we should make sure they have the same risk of dying in hospital due to comorbidities by controlling for them (i.e., both of them had uncomplicated diabetes), so that we can observe the true effect of obesity.

On the other hand, the most common method of controlling comorbidities is using comorbidities scores such as the Elixhauser score, which reflects the total number of comorbidities one patient has. This controlling method was used by most of the other studies exploring the associations between obesity and disease outcomes. In this example, both patients had only one disease comorbidity, and they would be considered at the same risk of dying in hospital if we used Elixhauser score (score=1). However, this may lead to the bias mentioned above. Only obese versus other patients with the same disease were compared (i.e., comorbidities controlled separately), should we discover the true effects of obesity on in-hospital mortality. In addition, eTable 4 suggested that the baseline characteristics were very different across the weight levels. As mentioned above, patients with obesity have lower numbers of Elixhauser scores than normal-weight patients (5.96 vs. 6.12). This result further supported that, patients with obesity had a lower number of comorbidities, but the diseases they presented tended to yield lower risks for in-hospital mortality. The disease trajectory, causality relationship, and patient management strategies for each comorbidity among patients with obesity warrant further investigation.

This study shares similar findings in population characteristics with other studies that explore the associations between obesity and disease outcomes. For example, Karagozian et. al. also used the adjusted regression model and found that age was associated with 2% higher odds of in-hospital mortality (OR=1.02, 95% CI: 1.01–1.03). Women were 14% less likely to die in hospital than men (OR=0.86, 95% CI: 0.75–1.00).^14^ Kwon et. al. found that compared to non-white patients, white patients were 22% more likely to die in the hospital after admission for stroke after adjustment for the baseline disease status (OR=1.22, 95% CI: 1.12, 1.33), which is similar to outcome findings that white patients had higher odds of in-hospital mortality than black patients.^15^

### Limitations

The study has several limitations. First, the severity of heart failure and whether it was with reduced or preserved ejection fraction were unknown, which might have a direct impact on mortality. Additionally, the normal BMI group was defined as BMI 20-24.9. This was limited by the ICD10 codes while clinically normal weight is defined as 18.8-25. This might lead to a small bias in the results. The reliability of the weight categories needs to be tested. However, it is consistent with other studies with similar methods. Second, this study, like other studies of hospitalized patients, is affected by selection bias caused by conditioning on the in-patient status, often called Berkson’s bias. For example, this study reported that black and Hispanic patients had lower risks of in-hospital death than white patients, which may be inconsistent with studies in outpatient settings. While others also reported similar findings, this may be due to the lower hospital access and utilization among black and Hispanic patients and those with more advanced diseases died outside of the hospital.

## Conclusions

Obesity is an independent risk factor for in-hospital mortality among patients admitted for heart failure. In-hospital mortality increases with the increase in BMI. Compared to normal-weight patients, obese patients have different comorbidity profiles in regard to the odds of in-hospital mortality, which may bias the outcome evaluation of heart failure treatment if these comorbidities were not considered separately.

## Data Availability

The data that support the findings of this study are available from Health Care Cost and Utilization Project (H-CUP) at AHRQ. Restrictions apply to the availability of these data, which were used under approval for this study. Data are available https://hcup-us.ahrq.gov/db/nation/nis/nisdbdocumentation.jsp with the permission of AHRQ.

## Appendix

**eTable 1.**
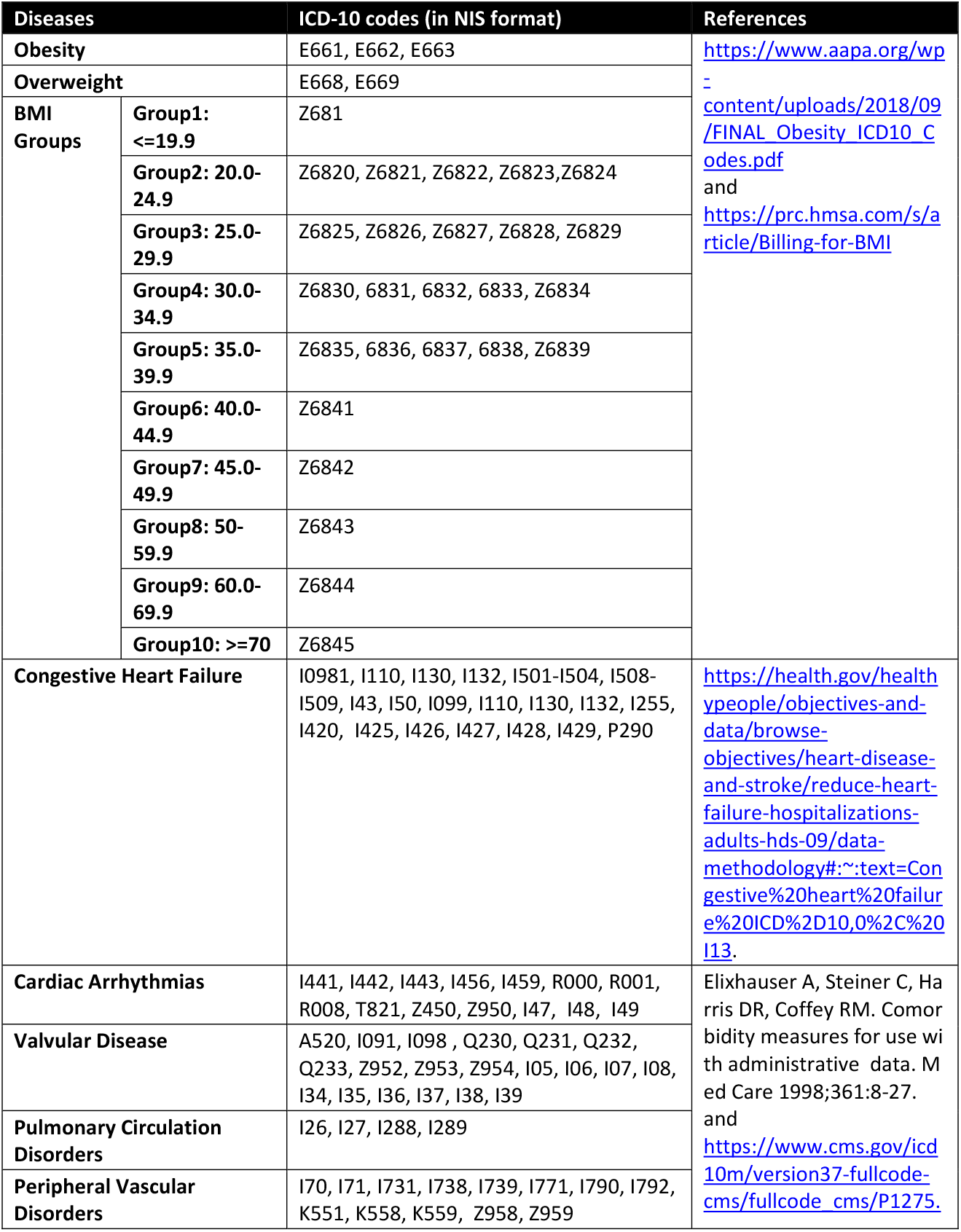
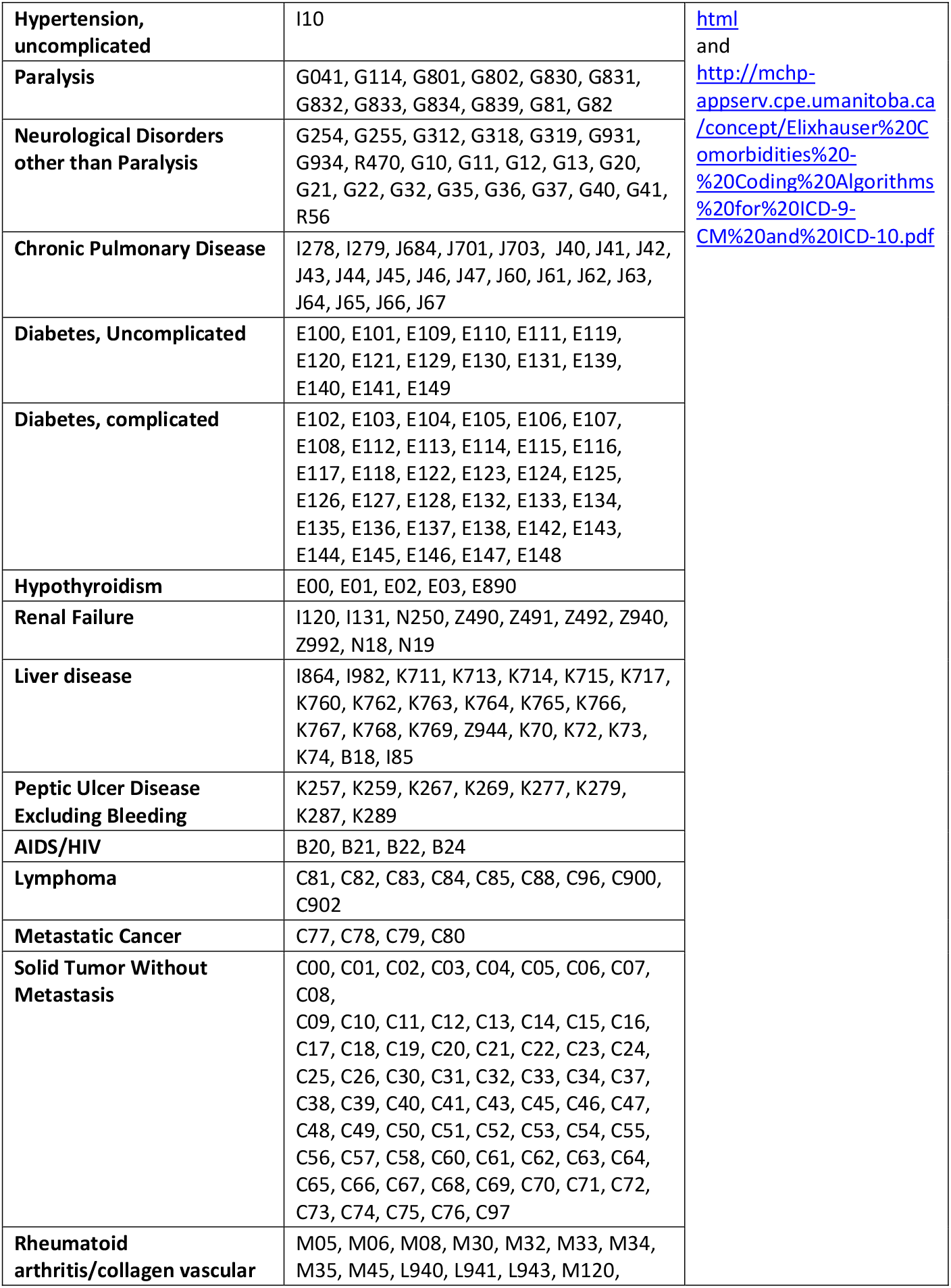
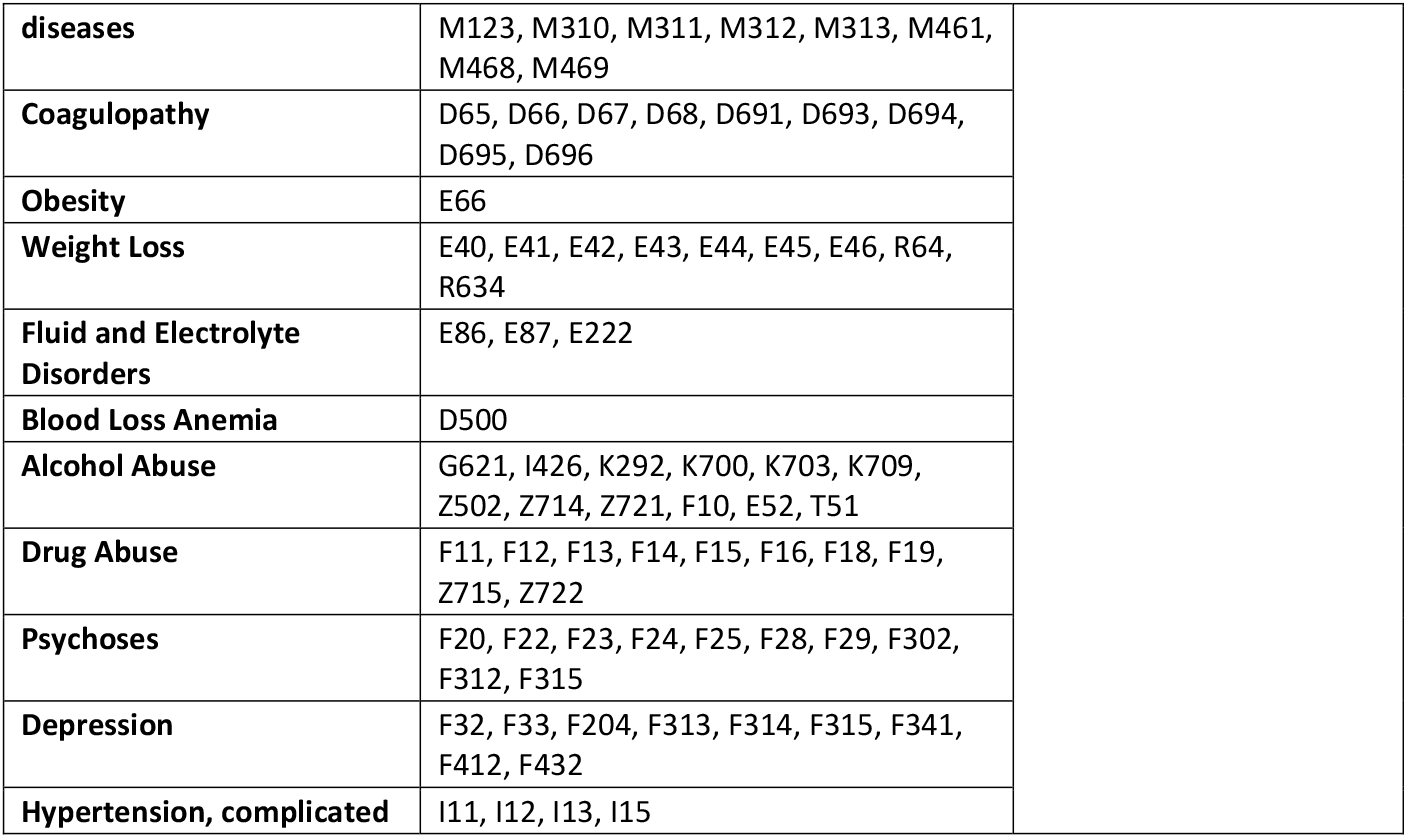
ICD10 codes used to identify study variables.

| Diseases |  | ICD-10 codes (in NIS format) | References |
| --- | --- | --- | --- |
| **Obesity** |  | E661, E662, E663 | https://www.aapa.org/wp-content/uploads/2018/09/FINAL_Obesity_ICD10_Codes.pdf<br>and<br>https://prc.hmsa.com/s/article/Billing-for-BMI |
| **Overweight** |  | E668, E669 |  |
| **BMI Groups** | **Group1: <=19.9** | Z681 |  |
|  | **Group2: 20.0-24.9** | Z6820, Z6821, Z6822, Z6823, Z6824 |  |
|  | **Group3: 25.0-29.9** | Z6825, Z6826, Z6827, Z6828, Z6829 |  |
|  | **Group4: 30.0-34.9** | Z6830, 6831, 6832, 6833, Z6834 |  |
|  | **Group5: 35.0-39.9** | Z6835, 6836, 6837, 6838, Z6839 |  |
|  | **Group6: 40.0-44.9** | Z6841 |  |
|  | **Group7: 45.0-49.9** | Z6842 |  |
|  | **Group8: 50-59.9** | Z6843 |  |
|  | **Group9: 60.0-69.9** | Z6844 |  |
|  | **Group10: >=70** | Z6845 |  |
| **Congestive Heart Failure** |  | I0981, I110, I130, I132, I501-I504, I508-I509, I43, I50, I099, I110, I130, I132, I255, I420, I425, I426, I427, I428, I429, P290 | https://health.gov/healthypeople/objectives-and-data/browse-objectives/heart-disease-and-stroke/reduce-heart-failure-hospitalizations-adults-hds-09/data-methodology#:~:text=Congestive%20heart%20failure%20ICD%2D10,0%2C%20I13. |
| **Cardiac Arrhythmias** |  | I441, I442, I443, I456, I459, R000, R001, R008, T821, Z450, Z950, I47, I48, I49 | Elixhauser A, Steiner C, Harris DR, Coffey RM. Comorbidity measures for use with administrative data. Med Care 1998;36:1:8-27.<br>and<br>https://www.cms.gov/icd10m/version37-fullcode-cms/fullcode_cms/P1275 |
| **Valvular Disease** |  | A520, I091, I098, Q230, Q231, Q232, Q233, Z952, Z953, Z954, I05, I06, I07, I08, I34, I35, I36, I37, I38, I39 |  |
| **Pulmonary Circulation Disorders** |  | I26, I27, I288, I289 |  |
| **Peripheral Vascular Disorders** |  | I70, I71, I731, I738, I739, I771, I790, I792, K551, K558, K559, Z958, Z959 |  |

|  |  |  |
| --- | --- | --- |
| **Hypertension, uncomplicated** | I10 | [html](http://mchp-appserv.cpe.umanitoba.ca/concept/Elixhauser%20Comorbidities%20Coding%20Algorithms%20for%20ICD-9-CM%20and%20ICD-10.pdf)<br>and<br>http://mchp-appserv.cpe.umanitoba.ca/concept/Elixhauser%20Comorbidities%20Coding%20Algorithms%20for%20ICD-9-CM%20and%20ICD-10.pdf |
| **Paralysis** | G041, G114, G801, G802, G830, G831, G832, G833, G834, G839, G81, G82 |  |
| **Neurological Disorders other than Paralysis** | G254, G255, G312, G318, G319, G931, G934, R470, G10, G11, G12, G13, G20, G21, G22, G32, G35, G36, G37, G40, G41, R56 |  |
| **Chronic Pulmonary Disease** | I278, I279, J684, J701, J703, J40, J41, J42, J43, J44, J45, J46, J47, J60, J61, J62, J63, J64, J65, J66, J67 |  |
| **Diabetes, Uncomplicated** | E100, E101, E109, E110, E111, E119, E120, E121, E129, E130, E131, E139, E140, E141, E149 |  |
| **Diabetes, complicated** | E102, E103, E104, E105, E106, E107, E108, E112, E113, E114, E115, E116, E117, E118, E122, E123, E124, E125, E126, E127, E128, E132, E133, E134, E135, E136, E137, E138, E142, E143, E144, E145, E146, E147, E148 |  |
| **Hypothyroidism** | E00, E01, E02, E03, E890 |  |
| **Renal Failure** | I120, I131, N250, Z490, Z491, Z492, Z940, Z992, N18, N19 |  |
| **Liver disease** | I864, I982, K711, K713, K714, K715, K717, K760, K762, K763, K764, K765, K766, K767, K768, K769, Z944, K70, K72, K73, K74, B18, I85 |  |
| **Peptic Ulcer Disease Excluding Bleeding** | K257, K259, K267, K269, K277, K279, K287, K289 |  |
| **AIDS/HIV** | B20, B21, B22, B24 |  |
| **Lymphoma** | C81, C82, C83, C84, C85, C88, C96, C900, C902 |  |
| **Metastatic Cancer** | C77, C78, C79, C80 |  |
| **Solid Tumor Without Metastasis** | C00, C01, C02, C03, C04, C05, C06, C07, C08, C09, C10, C11, C12, C13, C14, C15, C16, C17, C18, C19, C20, C21, C22, C23, C24, C25, C26, C30, C31, C32, C33, C34, C37, C38, C39, C40, C41, C43, C45, C46, C47, C48, C49, C50, C51, C52, C53, C54, C55, C56, C57, C58, C60, C61, C62, C63, C64, C65, C66, C67, C68, C69, C70, C71, C72, C73, C74, C75, C76, C97 |  |
| **Rheumatoid arthritis/collagen vascular** | M05, M06, M08, M30, M32, M33, M34, M35, M45, L940, L941, L943, M120, |  |

|  |  |
| --- | --- |
| **diseases** | M123, M310, M311, M312, M313, M461, M468, M469 |
| **Coagulopathy** | D65, D66, D67, D68, D691, D693, D694, D695, D696 |
| **Obesity** | E66 |
| **Weight Loss** | E40, E41, E42, E43, E44, E45, E46, R64, R634 |
| **Fluid and Electrolyte Disorders** | E86, E87, E222 |
| **Blood Loss Anemia** | D500 |
| **Alcohol Abuse** | G621, I426, K292, K700, K703, K709, Z502, Z714, Z721, F10, E52, T51 |
| **Drug Abuse** | F11, F12, F13, F14, F15, F16, F18, F19, Z715, Z722 |
| **Psychoses** | F20, F22, F23, F24, F25, F28, F29, F302, F312, F315 |
| **Depression** | F32, F33, F204, F313, F314, F315, F341, F412, F432 |
| **Hypertension, complicated** | I11, I12, I13, I15 |

**eTable 2.** The prevalence of comorbidities at the time of CHF admission (n=204,970)

| Comorbidities (Dx2-Dx40) | n | % |
| --- | --- | --- |
| Cardiac Arrhythmias | 109,758 | (53.55%) |
| Renal Failure | 107,566 | (52.48%) |
| Chronic Pulmonary Disease | 96,340 | (47.00%) |
| Diabetes, complicated | 93,721 | (45.72%) |
| Fluid and Electrolyte Disorders | 79,592 | (38.83%) |
| Pulmonary Circulation Disorders | 52,911 | (25.81%) |
| Valvular Disease | 49,396 | (24.10%) |
| Hypothyroidism | 37,133 | (18.12%) |
| Peripheral Vascular Disorders | 33,426 | (16.31%) |
| Depression | 30,304 | (14.78%) |
| Weight Loss | 29,641 | (14.46%) |
| Diabetes, uncomplicated | 28,749 | (14.03%) |
| Deficiency Anemia | 18,063 | (8.81%) |
| Neurological Disorders Other Than Paralysis | 16,168 | (7.89%) |
| Coagulopathy | 14,152 | (6.90%) |
| Liver disease | 11,207 | (5.47%) |
| Drug Abuse | 9,142 | (4.46%) |
| Rheumatoid arthritis | 7,111 | (3.47%) |
| Alcohol Abuse | 6,911 | (3.37%) |
| Solid Tumor Without Metastasis | 4,676 | (2.28%) |
| Hypertension, uncomplicated | 4,028 | (1.97%) |
| Psychoses | 2,829 | (1.38%) |
| Hypertension, complicated | 2,477 | (1.21%) |
| Metastatic Cancer | 1,911 | (0.93%) |
| Blood Loss Anemia | 1,892 | (0.92%) |
| Lymphoma | 1,653 | (0.81%) |
| Peptic Ulcer Disease Excluding Bleeding | 1,058 | (0.52%) |
| Paralysis | 1,001 | (0.49%) |
| AIDS/HIV | 332 | (0.16%) |

Note: CHF were excluded if reported Dx2-Dx40

**eTable 3.** Regression results of the associations between weight status without controlling for comorbidities

| Characteristics |  | Odds Ratio | 95% CI |
| --- | --- | --- | --- |
| Weight status | Normal | Reference |  |
|  | Underweight | 1.469* | (1.295 - 1.666) |
|  | Overweight | 0.742* | (0.642 - 0.858) |
|  | Obese Group 1 (BMI 30-34.9) | 0.433* | (0.379 - 0.496) |
|  | Obese Group 2 (BMI 35-39.9) | 0.428* | (0.374 - 0.491) |
|  | Obese Group 3 (BMI 40-44.9) | 0.584* | (0.513 - 0.664) |
|  | Obese Group 4 (BMI 45-49.9) | 0.584* | (0.504 - 0.678) |
|  | Obese Group 5 (BMI 50-59.9) | 0.563* | (0.480 - 0.660) |
|  | Obese Group 6 (BMI 60-69.9) | 0.636* | (0.503 - 0.805) |
|  | Obese Group 7 (BMI >= 70) | 1.077 | (0.824 - 1.408) |
| Age |  | 1.030* | (1.026 - 1.033) |
| Gender | Males | Reference |  |
|  | Females | 0.781* | (0.732 - 0.834) |
| Race | Whites | Reference |  |
|  | Blacks | 0.795* | (0.728 - 0.867) |
|  | Hispanics | 0.887 | (0.780 - 1.008) |
|  | Asian or Pacific Islander | 0.881 | (0.696 - 1.115) |
|  | Native American | 1.238 | (0.811 - 1.889) |
|  | Others | 1.003 | (0.803 - 1.252) |
| Length of Stay |  | 1.038* | (1.034 - 1.042) |
| Elixhauser Score |  | 1.268* | (1.246 - 1.290) |
| Year of Admission | 2017 | Reference |  |
|  | 2018 | 0.942 | (0.871 - 1.018) |
|  | 2019 | 0.930 | (0.861 - 1.004) |

\* p<0.05

**eTable 4.**
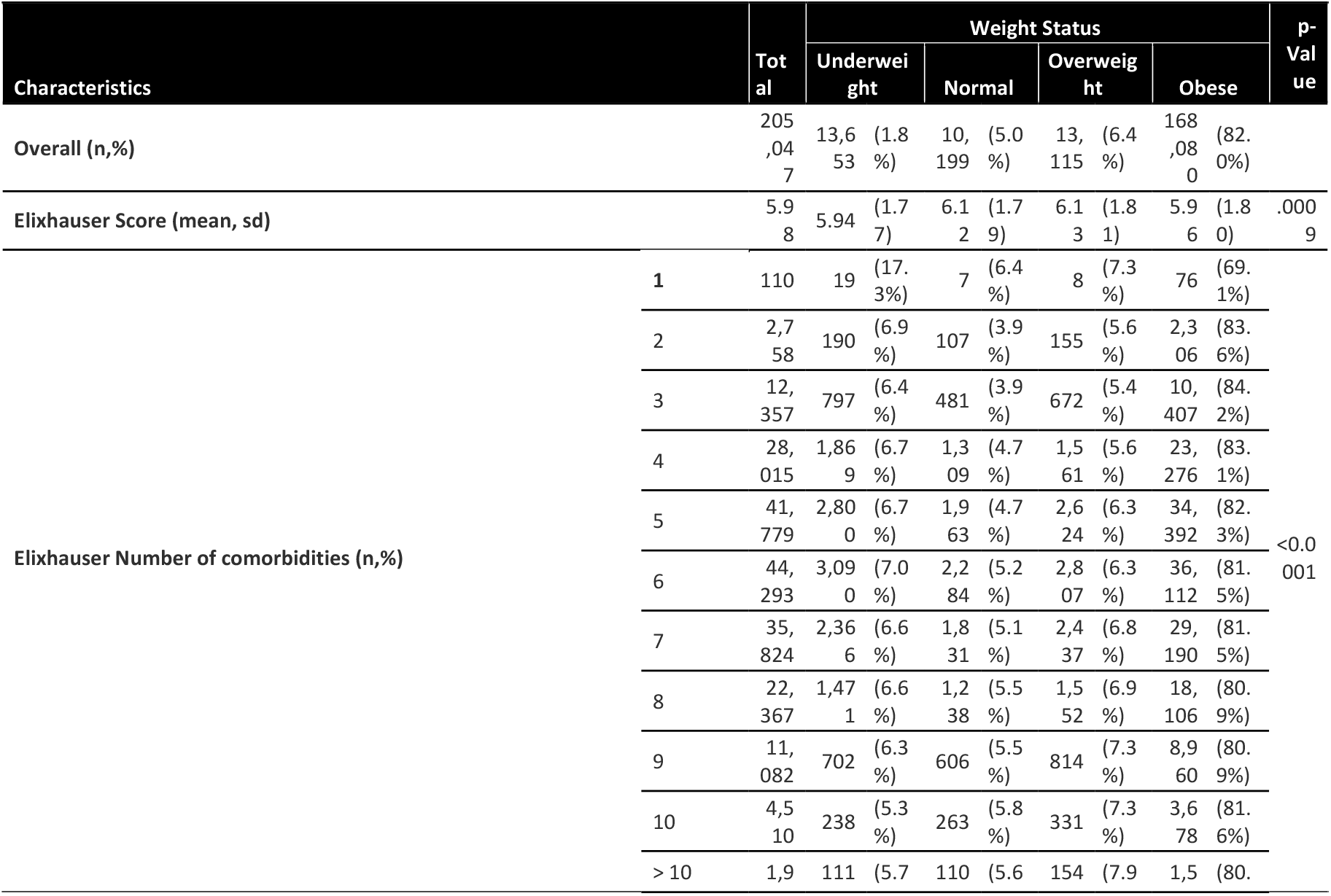
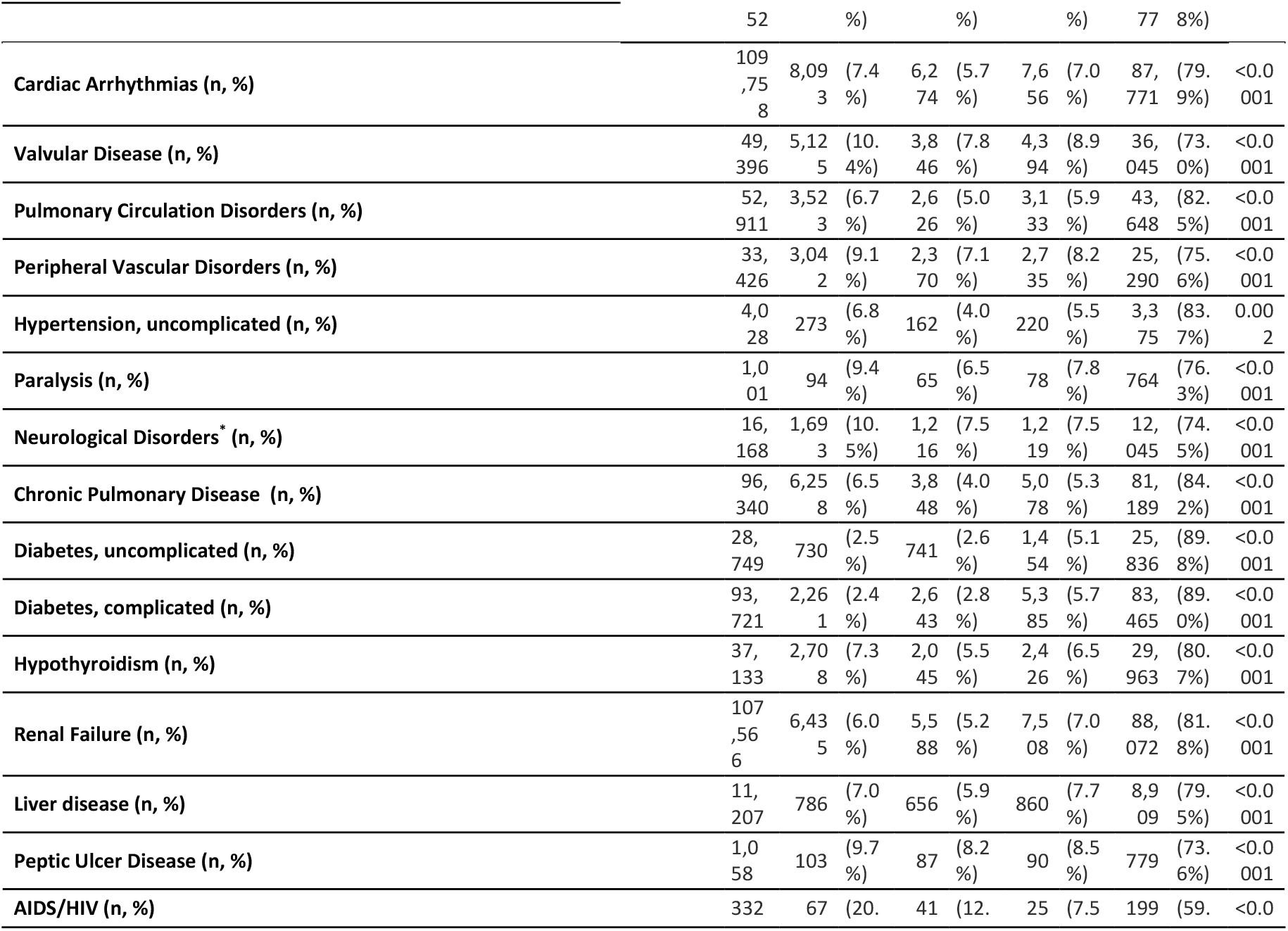
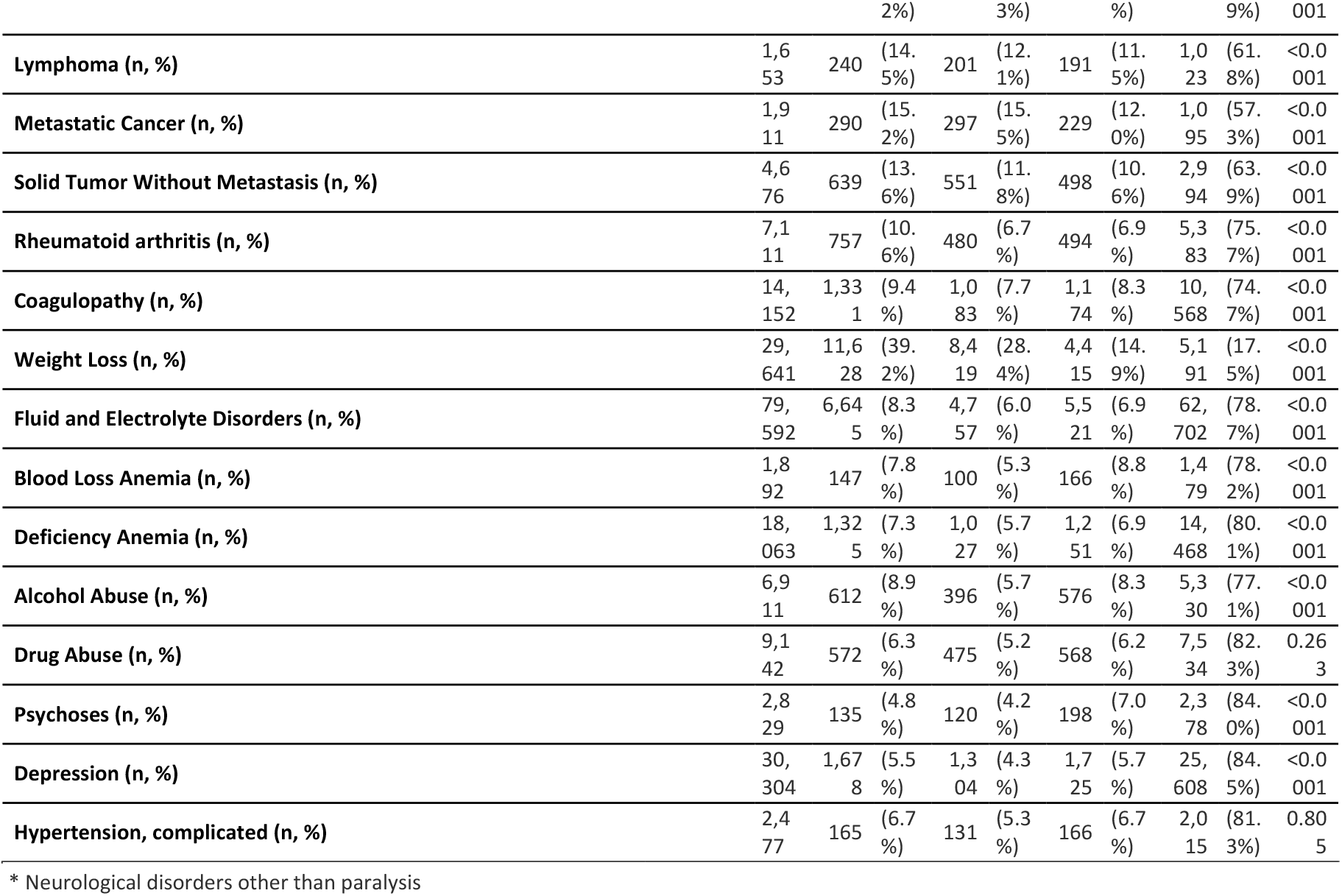
Baseline characteristics by weight status (descriptive and unadjusted)

## Notes

### Competing Interest Statement

The authors have declared no competing interest.

### Funding Statement

No external funding was received.

### Author Declarations

Health Care Cost and Utilization Project (H-CUP) at AHRQ approved usage of the data for the research. The data was de-identified.

